# Effects of lower tiers of government healthcare facilities on unmet need for contraception and contraception use in Bangladesh

**DOI:** 10.1101/2023.04.06.23288229

**Authors:** Md Nuruzzaman Khan, Shimlin Jahan Khanam, Md Mostaured Ali Khan, M Mofizul Islam, Melissa L. Harris

## Abstract

**Background:** In low- and middle-income countries (LMICs), including Bangladesh, modern contraception use remains lower than desired, resulting in a higher unmet need. A potential factor contributing to lower contraceptive use is reduced access to and use of lower tiers of government healthcare facilities, including home visits by family welfare assistants (FWAs), as well as women’s visits to community and satellite clinics. These relationships, however, are still unexplored in Bangladesh and LMICs more broadly. The aim of this study was to explore effects of lower tiers of government healthcare facilities on unmet need for contraception and contraception use in Bangladesh.

**Methods:** Data from 17,585 sexually active married women were analyzed from the 2017 Bangladesh Demographic and Health Survey. The outcome variables were any contraceptive use, modern contraceptive use, unmet need for contraception, and unmet need for modern contraception. The explanatory variables considered were respondent’s home visits by FWAs, respondent’s visits to a community clinic, and respondent’s visits to a satellite clinic. Multilevel mixed-effect Poisson regression with robust variance was used to determine the association between the outcome and explanatory variables, adjusted for individual-, household-, and community-level factors.

**Results:** Approximately 18% of respondents were visited by FWAs in the three months prior to the survey date and only 3.4% and 3.1% of women attended community and satellite clinics, respectively. Women who reported being visited by FWAs in the three months prior to the survey were approximately 36% less likely to report an unmet need for modern contraception and 42% more likely to report using modern contraception than women who did not report such a visit. A higher likelihood of unmet need for contraception and a lower likelihood of contraception use was found among women who did not visit these community or satellite clinics or visited these clinics for other reasons than collecting contraception as compared to women who visited these clinics to collect contraception.

**Conclusion:** FWAs’ visits to respondents’ homes to provide contraception, as well as respondent’s visits to satellite and community clinics play a major role in Bangladesh to ensure contraception use and reduce the unmet need for contraception. However, their coverage is quite low in Bangladesh. The findings suggest an urgent need for greater government initiatives to increase the number of FWAs and engage in proper monitoring them at the field level.

## Introduction

Every year, an estimated 121 million unintended pregnancies occur worldwide, accounting for 48% of the total 250 million pregnancies [1]. Around 92% (111.9 million) of these pregnancies occur in low- and middle-income countries (LMICs) - a percentage that is now increasing with the increasing number of reproductive aged (15-49 years) women in the total population [1, 2]. Nearly 61% of these pregnancies are ended by induced abortion. Access to safe abortion services however is restricted in LMICs with traditional or non-expert providers often the main source of abortion care despite limited knowledge of performing such care [1, 3-5]. This practice is responsible for approximately 192,000 maternal deaths every year in LMICs, and over 7 million women are admitted to hospital because of abortion related complications [6, 7]. Women who continue with unintended conceptions have also been shown to use maternal healthcare services less and engage in adverse health behaviours, which further increases adverse maternal and child health outcomes, including maternal and child mortality [2, 8-10]. Consequently, unintended conceptions are considered as ongoing public health threat in LMICs and a significant challenge to achieving the Sustainable Development Goal 3, which aims to ensure health and well-being for all by 2030 [2].

Key reasons for the higher prevalence of unintended pregnancy in LMICs is the non-use of contraception and an unmet need for modern contraception. This has been attributed to lower exposure to family planning messages, poor contraception knowledge and availability of emergency contraception [1, 2]. However, improving access to, and uptake of contraception in LMICs remains challenging, particularly for socio-economically disadvantaged women and those living in rural areas, despite the fact that the successful implementation of the Millennium Development Goals resulted in a significant increase in modern contraception use (from 52% to 62% in 2015) [11, 12]. Consequently, it has been found that around half of women of reproductive age who are in-union and living in LMICs like Bangladesh do not have proper access to modern contraception [13]. In Bangladesh, only 48% of women use modern contraception [14] and 15.5% have an unmet need for contraception, with these figures significantly higher in rural areas. Together, these behaviours, lead to a higher occurrence of unintended conception (48%) in the country [12].

Family planning services in most LMICs, including Bangladesh, play a critical role in providing access to, and use of, contraception [14, 15]. However, evidence suggests a significant decline in reliance on family planning services in Bangladesh in recent years, possibly due to the government’s focus on maternal and child health issues over family planning services [14]. As a result, there has been an increase in births resulting from contraception failure in Bangladesh in recent years [15].

Bangladesh has strong family planning services network countrywide through (i) Family Welfare Assistants’ (FWAs) visits to eligible couples’ (sexually active married women) homes every 14 days, (ii) providing access to contraception at the community level through 18,000 community clinics, and (iii) ensuring availability of contraception at satellite clinics [2]. Additionally, several non-governmental organizations (NGOs) provide family planning and contraception services, primarily in disadvantaged areas. Family planning and contraception services are also available in specialized private clinics and pharmacies; however, accessing these services requires a service charge. Therefore, their contributions in providing contraception to eligible couples is low. As a result, the government network continues to be the dominant source of contraception provision to eligible couples [2, 15]. However, little is known about the contribution of government sources of contraception provision, particularly with respect to the unmet need for contraception. It is critical for policymakers to identify which dimensions of family planning services are most effective now and where more emphasis should be placed to ensure universal access to contraception. However, this understanding is lacking in LMICs, including Bangladesh, and available studies have mainly examined socio-demographic factors associated with contraception use and the unmet need for contraception [16-21]. We therefore aimed to determine the effects of respondent’s home visits by FWAs, respondent’s visits to community clinics, and respondent’s visits to satellite clinics on contraception use and unmet need for contraception adjusted for potential confounders at the individual, household, and community-level.

## Methods

We analysed data from the 2017/18 Bangladesh Demographic and Health Survey, a nationally representative survey conducted every three years. The survey was administered by the National Institute of Population Research and Training, a subsidiary of the Ministry of Health and Family Welfare of Bangladesh. Financial and technical support was provided by several development partners, including USAID and UNFPA. The survey selected nationally representative households through a two-stage stratified random sampling method. At the first stage of sampling, the survey selected 675 enumeration areas (EAs, clusters) from the list of 293,579 EAs used by the Bangladesh Bureau of Statistics in the 2011 National Population Census. Of these EAs, data collection was undertaken in 672 EAs. Prior to data collection, a household listing operation was conducted. This was then used to select a fixed number of 30 households from each selected EA through probability proportional to the unit size. A total of 20,160 households were selected, of which data collection was undertaken in 19,457 households, resulting in a 96% inclusion rate. There were 20,376 eligible women aged 15-49 years who were usual residents of or lived in those households the night before the date of the survey. Data were collected from 20,127 women, resulting in a response rate of 98.8%. Details about this survey have been published elsewhere [14].

### Analytic sample

Data from 17,585 women who met the inclusion criteria were analysed in this study. The inclusion criteria were: (i) reproductive aged (15-49) married women with capacity to conceive, (ii) sexually active (at least one reported episode of sexual intercourse within one month preceding the survey), (iii) not currently pregnant and not in the post-partum amenorrhea period, (iv) not wanting to have a baby within 24 months of the interview date and (iv) responded to the questions related to contraception use and the unmet need for contraception.

### Outcome variables

We considered four outcome variables: (i) use of any contraception methods, (ii) use of modern contraception methods, (iii) unmet need for contraception, (iv) unmet need for modern contraception. These variables were based on the women’s responses to questions related to contraception use or non-use and their intention for future conception. At first, eligible women were asked, “*Are you or your husband currently doing something or using any method to delay or avoid getting pregnant*?” Responses were recorded dichotomously as Yes or No. Women who reported positively were then asked, “*Which method are you using*?” Where women reported multiple contraceptive methods, the most frequent contraceptive method was used. This response was then reclassified as any contraception use (Yes vs No) and modern contraception use (Yes vs No), using the World Health Organization’s recommended list of contraception [22]. Furthermore, women’s responses on contraception methods used were considered along with their intention for pregnancy and birth in the future to calculate the unmet need for contraception and unmet need for modern contraception. Data on future pregnancy intentions were collected by asking women, “*Would you like to have (a/another) child, or would you prefer not to have any (more) children*?” If the respondent wanted to have another child in the future, then they were asked, “*How long would you like to wait from now before the birth of (a/another) child*?” The variables unmet need for contraception (No vs Yes) and unmet need for modern contraception (No vs Yes) were generated if the respondent reported they did not want to have a baby in the future or near future (<24 months), but they were not using contraception or modern contraception, respectively.

### Study factors

The study variable was the accessibility of contraception methods. Women who reported using contraception were asked about the source of their last obtained contraception method. Given the majority of contraceptive services are provided by the government at the field level, only these sources were ascertained. Specifically, respondents were asked whether they had received the following services to collect contraception in the three months prior to the survey i) a community clinic (yes, no, respondent’s visited community clinic but did not collect contraception), ii) a satellite (yes, no, respondent’s visited satellite clinic but did not collect contraception), ii) a visit by FWAs (yes, no).

### Confounding variables

Potential confounding variables were identified by a comprehensive literature search [16-21]. The identified variables were women’s age (treated as a continuous variable), women’s educational status (no formal education, primary, secondary, and higher), women’s working status (yes, no), parity (no children, 1-2 children, >2 children), and intervals between the two most recent live births (≤2 years, 3-4 years, >4 years). Partner’s education (no formal education, primary, secondary, and higher), partner’s occupation (agriculture worker, physical worker, services, business, other), type of family (joint, nuclear), and wealth quintiles (poorest, poorer, middle, richer, and richest) were also considered. Other variables included were women’s place of residence (urban, rural), and region (Barisal, Chattogram, Dhaka, Khulna, Rajshahi, Mymensingh, and Sylhet).

### Statistical analysis

We explored the weighted distribution of the respondents’ socio-demographic characteristics through descriptive statistics, including frequency (percentage) and median (Q1-Q3). The effects of service accessibility on any contraception use, and modern contraception use, unmet need for contraception, unmet need for modern contraception were examined using a multilevel Poisson regression model. Separate models were run for each of the three outcome variables, with each model containing one explanatory variable: respondent’s home visits by FWAs, respondent’s visits to community clinics, and respondent’s visits to satellite clinics, along with other individual, household, and community-level factors. We used a multilevel Poisson regression model because the survey data we analyzed were clustered. This is important to obtain accurate results particularly when prevalence of the outcome variables is higher than 10%. Previous studies have shown that the logistic regression model overestimates the true likelihood if the prevalence of the outcome variable is higher than 10%. The multilevel Poisson regression model can consider both the clustering structure of the data and address the issue of overestimation. We checked for multicollinearity before including variables in the models. If evidence of multicollinearity was found, we deleted the relevant variable and ran the model again. Results were reported as Prevalence Ratios (PR) along with their 95% Confidence Intervals (95% CI). All statistical analyses were conducted using Stata software version 15.1 (Stata Corp, College Station, Texas, USA).

## Results

### Background characteristics of the respondents

The weighted percentage of the respondents’ socio-demographic characteristics are represented in Table 1. The median age of the respondents was 32 years. Around 38% of the total respondents had secondary level education. Almost 6% of respondents had not given birth at the time of the survey and among the women, who had children, around 11% had a birth interval of less than 2 years. Nearly 20% of the interviewed sample were from poorest quintile and 72% resided in rural areas.

**Table 1:** Background characteristics of the respondents.

| Characteristics | n (%) |
| --- | --- |
| Women's age, median (Q1-Q3) | 32 (25-40) |
| **Women's education status** |  |
| No formal education | 3157 (17.9) |
| Primary | 5712 (32.5) |
| Secondary | 6760 (38.4) |
| Higher | 1956 (11.2) |
| **Women's employment status** |  |
| Yes | 8986 (51.1) |
| No | 8599 (48.9) |
| **Partner's education status** |  |
| No formal education | 3772 (22.9) |
| Primary | 5322 (32.4) |
| Secondary | 4800 (29.2) |
| Higher | 2510 (15.3) |
| **Partners' occupation** |  |
| Agriculture worker | 4424 (27.6) |
| Physical worker | 7380 (46.0) |
| Services | 837 (5.2) |
| Business | 3361 (20.9) |
| Other | 38 (0.28) |
| **Parity** |  |
| No children | 1091 (6.2) |
| 1-2 children | 8610 (49.0) |
| >2 children | 7884 (44.8) |
| **Intervals between the two most recent live births** |  |
| ≤2 years | 1974 (11.2) |
| 3-4 years | 4219 (24.0) |
| >4 years | 11392 (64.8) |
| **Family type** |  |
| Nuclear family | 7614 (43.3) |
| Joint family | 9971 (56.7) |
| **Household wealth status** |  |
| Poorest | 3356 (19.1) |
| Poorer | 3537 (20.1) |
| Middle | 3560 (20.3) |
| Richer | 3621 (20.6) |
| Richest | 3511 (20.0) |
| **Place of residence** |  |
| Urban | 4908 (27.9) |
| Rural | 12677 (72.1) |
| **Region (administrative division)** |  |
| Barishal | 1005 (5.7) |
| Chattogram | 3164 (18.0) |
| Dhaka | 4381 (24.9) |
| Khulna | 2074 (11.8) |
| Mymensingh | 1323 (7.5) |
| Rajshahi | 2472 (14.1) |
| Rangpur | 2129 (12.1) |
| Sylhet | 1036 (5.9) |

#### Prevalence of contraception use and unmet need for modern contraception

The percentage of individuals using any form of contraception was 63.1%, while the percentage of those using modern contraception was 52.9% (Table 2). Additionally, the reported percentage of unmet need for contraception was 13.5%, and the unmet need for modern contraception was 24.4%.

**Table 2:**
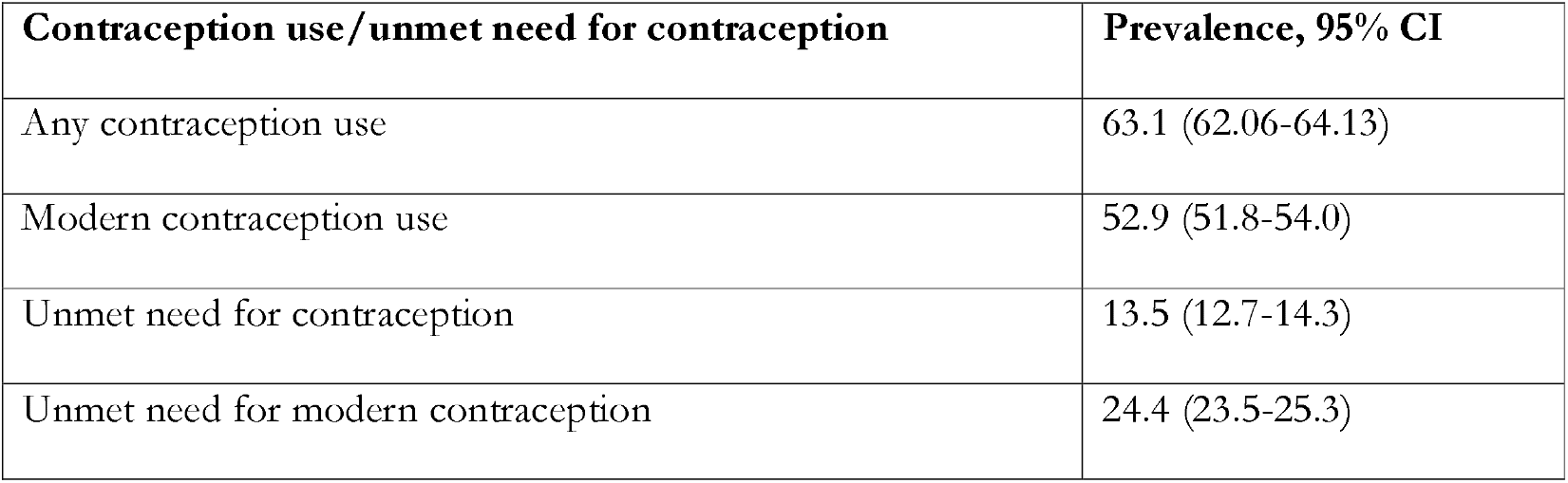
Distribution of contraception use and unmet need for contraception, Bangladesh, 2017/18.

| Contraception use/unmet need for contraception | Prevalence, 95% CI |
| --- | --- |
| Any contraception use | 63.1 (62.06-64.13) |
| Modern contraception use | 52.9 (51.8-54.0) |
| Unmet need for contraception | 13.5 (12.7-14.3) |
| Unmet need for modern contraception | 24.4 (23.5-25.3) |

**Figure 1:** Source of collecting contraception across respondents’ wealth quintile

#### Access to lower tiers of government healthcare facilities for obtaining contraception

The distribution of the respondents based on the sources of obtaining last form of modern contraception are presented in Table 3. FWAs provided last form of modern contraception to 18.1% of the total respondents. Around 3.4% and 3.1% of respondents reported they obtained their last form of modern contraception from satellite and community clinics, respectively. Together, around 23% of respondents reported that they collected their last form of modern contraception from one of the three sources in the three months prior to the survey.

**Table 3:**
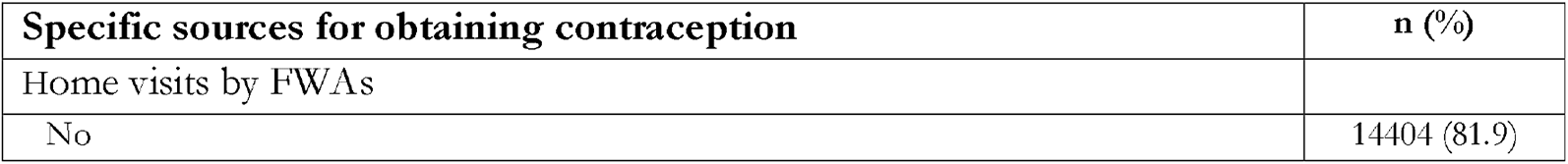
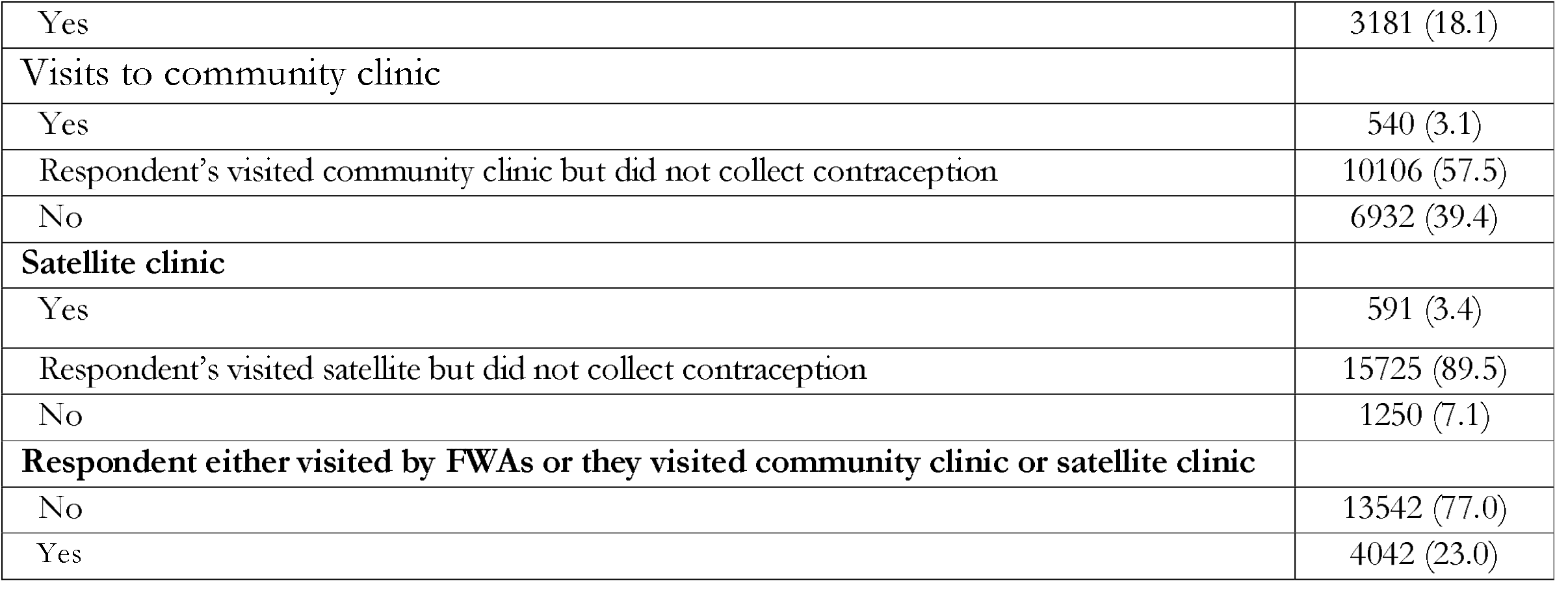
**Distribution of specific sources for obtaining contraception (**Family welfare assistants, **community clinic, and satellite clinic) in Bangladesh in the three months prior to the survey completion, BDHS, 2017/2018**.

#### Distribution of contraception use and unmet need for contraception across respondents’ contraception accessing options

The distribution of use of any contraception use, and modern contraception use, unmet need for contraception, and unmet need for modern contraception across respondents’ source of contraception are presented in Figure 1. The use of any contraception (84.47% vs 56.72%) and modern contraception (78.03% <u>vs</u> 45.42%) were found significantly higher among women who either visited community or satellite clinics or FWAs visited their homes to provide contraception. We found unmet need for contraception (6.9% vs 15.63%) and unmet need for modern contraception (13.38% vs 27.94%) were around 50% lower among the respondents collected contraception from community clinic or satellite clinic or visited by FWAs to provide contraception.

**Figure 1:**
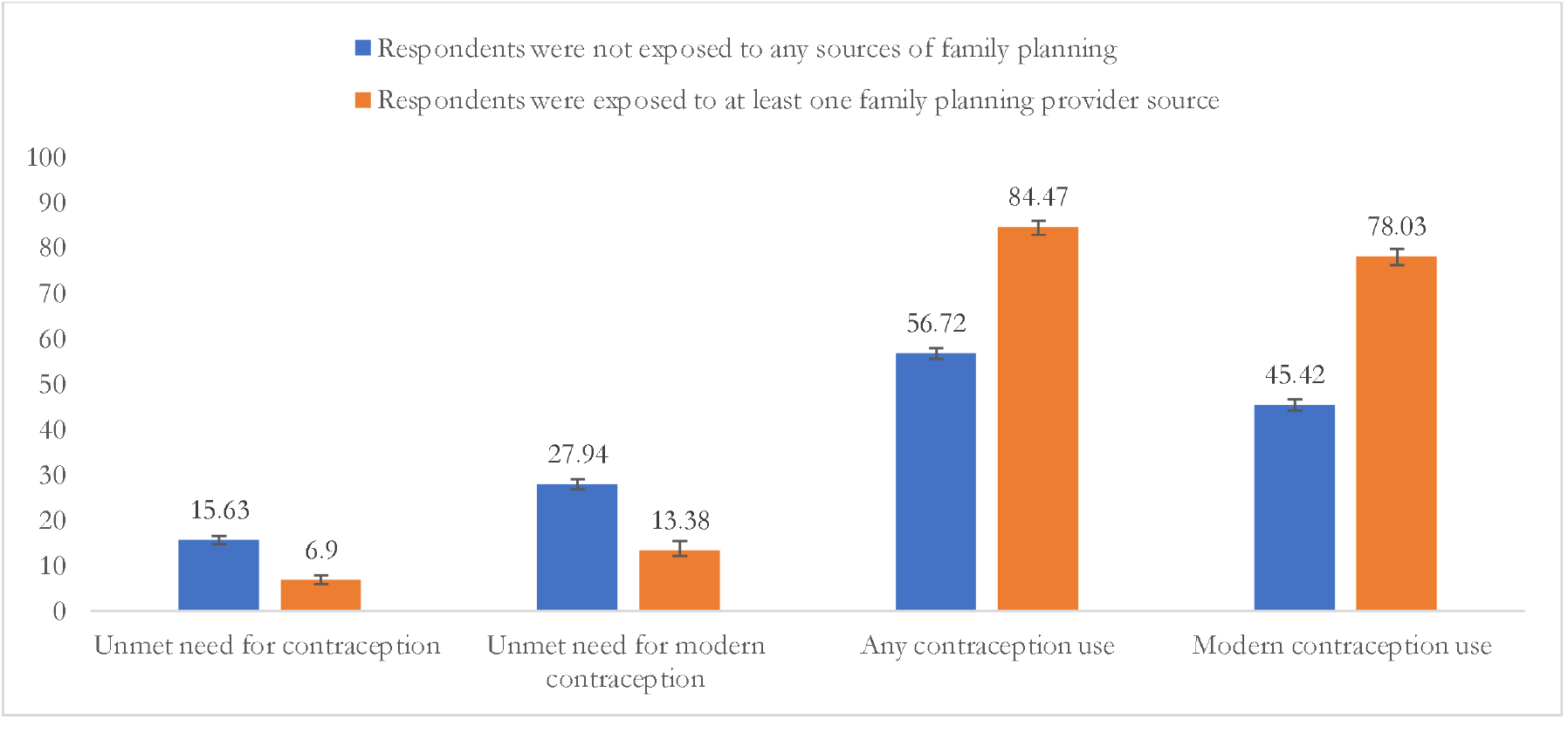
Contraception use and unmet need for contraception use status across respondents’ contraception accessing options

#### Lower tiers of government healthcare facilities and its association with contraception use and unmet need for contraception

The effects of contraception accessing options on contraception use and unmet need for contraception are presented in Table 4. A 36% lower prevalence of unmet need for modern contraception (aOR, 0.64, 95% CI, 0.58-0.70) and 42% lower prevalence of unmet need for contraception (aOR, 0.58, 95% CI, 0.51-0.67) among women who were visited by FWAs than those who were not was found. In addition, around 3 to 11 times higher likelihoods of unmet need for contraception and unmet need for modern contraception were found among women who did not obtain their contraception from either a community clinic or satellite clinic or visited community clinic or satellite clinic but did not collect contraception as compared to the women who collected contraception from the community clinic or satellite clinic. Having at least one visit by FWAs in the three months prior to the survey date to provide contraception was associated with a 1.34 (95% CI, 1.31-1.38) times higher likelihood of using any contraception method and a 1.46 (95% CI, 1.41-1.51) times higher likelihood of modern contraception use. Around 31%-46% lower likelihoods of any contraception use and modern contraception use was found among women who reported either visits to community or satellite clinics, but not to collect contraception, as compared to the respondents who reported they collected contraception from any of these sources. Exposure to at least one of the three sources resulted in 51-54% lower likelihoods of unmet need for contraception and 1.46-1.66 times higher likelihoods of any contraception use or modern contraception use. We also conducted separate analyses for rural and urban samples to check whether lower tiers of government healthcare facilities operate differently in rural and urban areas. However, we did not report any significant differences (results not shown in the table).

**Table 4:** **Government sources of obtaining contraception** in the three months prior to the survey **and its association with unmet need for contraception and contraception use**

| Service | Any contraception use, PR (95% CI) | Modern contraception use, PR (95% CI) | Unmet need for contraception, PR (95% CI) | Unmet need for modern contraception, PR (95% CI) |
| --- | --- | --- | --- | --- |
| Respondent's home visits by FWAs |  |  |  |  |
| No | 1.00 | 1.00 | 1.00 | 1.00 |
| Yes | 1.34 (1.31-1.38) *** | 1.46 (1.41-1.51) *** | 0.58 (0.51-0.67) *** | 0.64 (0.58-0.70) *** |
| **Community clinic** |  |  |  |  |
| Respondent's visits to community clinic | 1.00 | 1.00 | 1.00 | 1.00 |
| Yes | 0.64 (0.62-0.66) *** | 0.54 (0.52-0.56) *** | 10.62 (5.09-22.15) ** | 12.15 (6.81-21.67) ** |
| Respondent's visited community clinic but did not collect contraception | 0.66 (0.64-0.69) *** | 0.57 (0.55-0.60) *** | 9.12 (4.38-18.99) ** | 10.82 (6.09-19.24) ** |
| **Satellite clinic** |  |  |  |  |
| Yes | 1.00 | 1.00 | 1.00 | 1.00 |
| Respondent's visited satellite but did not collect contraception | 0.67 (0.65-0.69) *** | 0.57 (0.55-0.59) *** | 4.27 (2.67-6.83) *** | 5.59 (3.69-8.48) ** |
| No | 0.69 (0.65-0.73) *** | 0.59 (0.55-0.64) *** | 3.55 (2.17-5.81) *** | 4.95 (3.22-7.60) *** |
| **Respondent either visited by FWAs or they visited community clinic or satellite clinic** |  |  |  |  |
| No |  |  | 1 |  |
| Yes | 1.46 (1.42-1.50) *** | 1.66 (1.61-1.72) *** | 0.46 (0.40-0.54) *** | 0.49 (0.45-0.55) *** |

Note: Models are adjusted for women's age, women' education, women's working status, partner's education, partner's occupation, parity, interval between two most recent life birth, family types, wealth quintiles, place of residence and place of region.
\*\*\*p<0.01, \*\*p<0.05

## Discussion

In this study, we investigated the effectiveness of lower tiers of government healthcare facilities, such as, respondent’s home visits by FWAs, respondent’s visits to community clinic, and respondent’s visits to satellite clinic on the use of contraception and unmet needs for contraception, while adjusting for individual, household, and community-level factors. We found that approximately 23% of all reproductive-aged women in Bangladesh obtained contraception from one of these three sources in the three months prior to the survey, with the majority receiving it from FWAs. After accounting for various factors, our fully adjusted model revealed that access to each of the three sources was a significant predictor of contraception use and non-access increased the likelihood of unmet needs for contraception. These findings underscore the importance of strengthening the government’s family planning options at the field level to promote the use of modern contraception and decrease unmet needs for contraception in Bangladesh. This could aid Bangladesh in achieving its SDGs targets by ensuring universal access to sexual and reproductive healthcare services and reducing maternal and under-five mortality rates.

Bangladesh has achieved historic progress in expanding access to contraception use over the last 50 years with around 54% of the total eligible population now using modern contraception compared to 7% in 1974 [15]. This historic progress is largely due to the government efforts that began in 1972 by initiating an independent family planning division under the Ministry of Health and Family Welfare that has invested in and delivered major policies and programs in the subsequent years [23]. These include a declaration of rapid population growth as the number 1 problem in Bangladesh and its focus on the national population policy outline (1975-1980), formation of upazila (second administrative unit of Bangladesh) family planning committee (to monitoring family planning activity) (1980-1985), formation of satellite clinics (to ensure family planning availability in remote and rural area) and initiation of unit-wise FWA registers (to keep family planning and other demographic record) (1985-1990) and consideration of family planning programs in subsequent health, nutrition and population sector program (1998-present) [2]. However, Bangladesh’s family planning agenda remains incomplete with very slow progress since the early 2000’s - a time when non-use of modern contraception and unmet need for modern contraception were 50% and 27%, respectively. Research has shown that that this situation remains mostly unchanged [15]. The reasons for such stagnation are many, however, lack of sustainability of the policies and programs and reducing focus on family planning to prioritise other maternal and child health indicators as targeted in the Millennium Development Goals in 2000 have been suggested as core reasons [2, 15].

One of Bangladesh’s most successful family planning initiatives involved FWAs visiting eligible women’s homes every 14 days to provide contraception counselling and supplies, as well as the use of mass media to raise awareness about family planning [24]. In 2000, approximately 46% of eligible women received FWAs’ visits and 50% learned about contraception through mass media, with even higher contributions among rural and disadvantaged populations [24, 25]. However, despite its reported success, FWAs’ visits have declined over time, with only 18% of respondents reporting such visits in this study. This reduction in FWAs’ visits is concerning given their past success in increasing contraception uptake and reducing unmet need for contraception, and several challenges at the respondent, provider, and policy levels may be contributing to this decline.

Bangladesh has made significant progress in women’s education and their participation in formal work, means less availability for at home services compared to the 2000s [26]. Consequently, the previous approach of providing family planning and contraception through FWAs visiting women’s homes is no longer effective for many women. The main reason is that the visits are made during the day when women are at work, making it difficult for them to receive information and methods for contraception, even if they have such intention [15, 27]. In addition, there is a shortage of FWAs, with approximately 40% of FWAs’ positions currently vacant [27]. The government has not been successful in addressing this issue due to structural challenges, including current vacancies of FWAs and educational requirements to undertake FWA roles. The minimum educational qualification required for FWAs recruitment is higher secondary level education, with no restriction on the maximum educational requirement. However, the flexible criteria for the maximum educational level required, combined with the current higher unemployment rate in Bangladesh among higher educated people, means that most FWAs have graduate or post-graduate degrees. They often continue to search for better jobs after working as an FWAs because the position is considered a low-status government job in Bangladesh, and the salary offer is much lower than other jobs with similar qualifications. Often, they leave the current position as FWAs once they find another better job, making it vacant. Furthermore, the current number of FWAs positions is inadequate, as the positions were created in the 1980s when the population of Bangladesh was around half of what it is now. This has led to reduced service quality, fewer visits, and a reduction in contraception use, resulting in an increase in unmet need for contraception.

The challenges encountered by the FWAs are also responsible for the declining roles of community clinics and satellite clinics in the provision of contraception [28]. In this study, we found that community clinics and satellite clinics play an important role in increasing contraception uptake and reducing unmet needs. The underlying reason is that FWAs are mostly responsible for running satellite clinics, and their absence or lack of presence leads to the unavailability of satellite clinics. Even if the satellite clinics are available, a lack of FWAs may lead to inadequate focus on family planning and contraception because child vaccinations are given higher priority than family planning [25]. There are around 18,000 community clinics in Bangladesh run by the government’s recruited MBBS doctors, but it also faces similar challenges like the satellite clinics. For instance, MBBS doctors do not feel comfortable staying in the village due to the lack of accommodation, transportation, and other residential facilities. Consequently, community clinics mostly run without the presence of healthcare personnel, or limited healthcare personnel [28]. This irregularity of healthcare provision leads to other healthcare taking precedence (e.g., district level hospital). In addition, patients also do not feel comfortable accessing family planning services and contraception with others present, as it is a culturally sensitive issue. Since 2010, when family planning and contraception became available in upazila and district-level hospitals, healthcare personnel in community clinics may have thought that respondents should collect contraception from these sources, although, in reality, this is rare at the field level [2]. Along with these challenges, the current achievement of replacement-level fertility should motivate the government to reduce the focus on family planning and contraception at the field level and give more focus on population management [27]. However, whatever the challenges or intentions are, it is important to ensure that family planning and contraception are easily accessible at government-supported facilities to increase contraception use and reduce unmet needs, leading to the improvement of maternal and child health.

The implications of our findings are that the government-supported family planning and contraception options, including FWAs visits, satellite clinics, and community clinics, continue to play a major role in increasing contraception uptake and reducing unmet need. Therefore, there needs to be substantial investment to strengthen these options and improve their ability to provide family planning and contraception services. It is also important to ensure an adequate number of FWAs at the field level by creating new FWAs’ posts and filling them, as well as providing them with better salaries and other opportunities for a better lifestyle. Separate family planning and contraception facilities should be established in all satellite clinics and community clinics, along with proper monitoring at all levels.

The current study has several strengths and a few limitations. This is the first study of its kind that provides evidence of the importance of sustaining FWAs’ visits, satellite clinics, and community clinics at the community level at a time when the focus on all three levels is declining. The data for this study was also extracted from a nationally representative survey and analyzed through advanced statistical methods with a range of individual, household, and community-level factors. Therefore, the findings of this study are more precise and should be used for national-level policy and program-making. However, the data analyzed in this study was cross-sectional in nature, and the reported associations are therefore correlational only and not causal. The data was also collected retrospectively, and respondents were asked to answer on past events; therefore, there may be recall bias in the responses. However, to reduce this bias, respondents were asked several follow-up questions on their past events, and any biases are likely to be random. In addition, besides these three avenues of providing family planning and contraceptives by the government, there are also upazila (second administrative unit of Bangladesh) to tertiary level hospitals. Also, anyone can obtain contraception from local pharmacies and private facilities. However, these data are not available in the survey, so they were not considered in this study. Despite these limitations, the associations reported in this study provide an important direction about the massive role that these three-family planning and contraception providing options of the government at the field level play to ensure contraception use and reduce unmet need.

## Conclusion

In this study, modern contraceptive use was found to be low with only half of women using these methods. In addition, a quarter of women were found to have an unmet need for contraception.

FWAs’ visits to respondents’ homes, as well as visits to community clinics and satellite clinics, were found to play an important role in increasing modern contraceptive uptake and reducing unmet need for modern contraception. These findings suggest the need for policies and programs to strengthen these government-based family planning and contraception services at the field level to reduce the current high rates of contraception non-use and unmet need. A core part of these strategies will be to address the existing lack of FWAs at the field level by identifying ways to educate, recruit and retain these vital workers.

## Data Availability

All data produced are available online at DHS data archive

https://dhsprogram.com/data/

## Acknowledgement

The authors thank the MEASURE DHS for granting access to the 2011 and 2017/18 BDHS data.

## Authors’ contributions

MNK designed the study, performed the data analysis, and wrote the first draft of this manuscript. SJK, MAK, MMI and MLH critically reviewed and edited the previous versions of this manuscript. All authors approved this final version of the manuscript.

## Data availability

The datasets used and analysed in this study are available from the Measure DHS website: https://dhsprogram.com/data/available-datasets.cfm

